# Establishing attributes and corresponding levels for a discrete choice experiment on nurses’ job preferences in Iran

**DOI:** 10.1101/2022.10.18.22281235

**Authors:** Sara Emamgholipour, Mahdi Shahraki, Amir Mohammad Haddadfar

## Abstract

**Background:** DCE is a widely used technique in healthcare to elicit non-market-stated preferences. This study was conducted primarily to identify the most important factors affecting the job preferences of nurses through a scoping review of literature review and qualitative methods, and finally to help select attributes and attribute levels.

**Methods:** This study was conducted in four stages based on Helter and Boehler’s proposed framework. The first stage was raw data collection to identify the factors influencing nurses’ job preferences. For this reason, a scoping review was performed to identify relevant attributes and their levels. In addition, 7 nurses and experts in the field of nursing were interviewed (in-depth interviews) to identify related characteristics according to the Iranian nursing workplace. Then, 19 nurses were asked to rate the attributes and levels. At this stage, all possible attributes from different sources were gathered. In the third stage, inappropriate attributes were deleted based on acquired qualitative data, and the research team decided which attributes to include in the final design. Attribute wording was done in the final stage. JMP Pro 16 was used to construct the final design. A utility-neutral design was generated and blocked into 3 versions, each consisting of 6 scenarios (choice sets). Finally, a pilot study involving 24 nurses was undertaken in April 2022.

**Results:** By using scoping review and qualitative methods such as semi-structured interviews, 23 potential job-related factors that affect nurses’ career choices were identified. Salary, type of employment contract, workload, type of health facility, workplace facilities, work schedule, and Expected time spent on the assigned job for promotion to a higher position were 7 attributes considered in the final design of the scenarios. Internal and face validity, intelligibility, and acceptability of the questionnaire were tested in the pilot study, and minor modifications were made. It was revealed that the respondents in the pilot study were able to understand and answer all of the choice sets with ease.

**Conclusion:** Some of the most significant financial and non-financial factors that affect Iranian nurses’ job preferences are mentioned in this study. This indicates that policymakers have a wide range of interventions available that can significantly improve the working conditions of nurses. Additionally, the full description of the attribute development and level selection processes makes this study valuable to the literature on DCE.

## Introduction

The decline in the number of healthcare practitioners is one of the most important concerns in today’ s healthcare system. Furthermore, the inequality in healthcare worker distribution between large metropolitan cities and remote or non-capital areas has become a serious concern and a top priority to address (1,2). Even though it is a global problem (3,4), low and middle-income countries have made significant efforts to provide the best possible response to the topic of how to recruit and retain healthcare workers in neglected areas (5). Additionally, from the perspective of the healthcare recipient who lives in rural locations or is distant from hospitals, there is a strong desire to get high-quality medical treatment from knowledgeable and competent healthcare professionals at a reasonable cost (6).

The nursing staff is a critical component of improving the healthcare system and delivering high-quality, effective, and convenient care. The shortage and mal-distribution of nursing staff are partly responsible for the increasing workload of existing nurses, which may decrease the quality of health services and patient satisfaction, and even cause conflict between nurses and patients (7).

Information on health worker preferences for job attributes can help guide policymakers in deciding which strategies to pursue to attract essential health workers to underserved areas. Discrete choice experiments are a feasible policy tool that can be used in developing countries to elicit such preference information (8).

DCE is a quantitative method that incorporates random utility theory (9), consumer theory (10), experimental design theory (11), and econometric analysis (12). DCEs have become more commonly used in healthcare settings, primarily to value patient experiences and evaluate trade-offs between health outcomes and patient experiences (13). DCE has also been used in the field of human resources for health (HRH), specifically to find incentives to attract and/or retain healthcare personnel (14). It is a well-suited method to elicit the stated job preferences of health workers, given specified attributes and levels (15,16). DCE investigations can reveal which job qualities are more important and which are less important from the healthcare worker’ s perspective. The policy importance of the resulting preferences may be determined not only by how strong the specific options are but also by how realistic they are from the views of policymakers and health workers, as well as by the labor market’ s context-specific characteristics (14).

Despite its widespread use around the world, DCE has only been used in a few studies in the health sector in Iran (17–21) One study assessed medical students’ preferences for working in deprived areas in Kermanshah (a province located in the west of Iran) (22). There hasn’ t been any research done in Iran on the career choices of nurses.

## Methods

The DCE is based on the concept that any product, service, or intervention can be described by its attributes, and that the value of the product, service, or intervention is determined by the levels of the attributes (23). Hence, the first stage in this process is to choose attributes and levels that can be used to characterize the product or service. This is one of the most important elements in the study design process (24). There is no single way to select attributes and levels, and it can be obtained through one of several methods. Multiple approaches, such as literature review, focus group discussion (FGD), interview, and experts’ panel, or a mix of these methods, can be used to identify the attributes and levels (23,25). Additionally, applying qualitative methods to elicit opinions and perspectives from possible beneficiaries is a recommended method for determining attributes and levels (24,26). This study aimed to identify factors related to nurses’ job preferences by carrying out a scoping review and conducting interviews with nurses and experts in nursing. Subsequently, establishing and preparing the most appropriate attributes and corresponding levels for further research.

### Conceptual framework

A framework proposed by Helter and Boehler was used in this study (27). This framework has been also used in other studies to systematically develop attributes and levels for conducting DCEs (28). In this framework, the researchers provide a systematic approach to attribute development for health-related DCEs and recommend following a four-stage process consisting of raw data collection, data reduction, removing inappropriate attributes, and wording of attributes (28). First, raw data about attributes and levels are collected using qualitative studies and alternative methods such as literature reviews. Then, the collected data are reduced through analysis. This results in a long list of attributes and levels. Moreover, these are then screened for appropriateness considering multiple criteria to reduce them to a limited number of attributes and levels. Finally, the attributes and levels are worded using methods such as piloting, and researchers’ judgment (28).

#### Stage 1: Data collection

This study was carried out in four stages. The first stage was raw data collection. This stage involves identifying the attributes relevant to the stated research question and then assigning levels for each of these attributes (29,30). This is an important component of the design since these attributes and attribute levels reflect the hypothetical circumstances considered in the DCE. Therefore, the study’ s underlying validity depends on the researcher’ s ability to correctly specify the relevant attributes (29). Thus, to come up with a main list of attributes, a scoping review was conducted. A mixture of regularly used keywords linked to the topic of conjoint analysis and the job preferences of healthcare workers were searched in the PubMed, Scopus, Google Scholar, and Web of Science databases. The following keywords were used in the search: discrete choice experiment, conjoint analysis, conjoint choice model, discrete choice model, conjoint choice experiment, stated preferences, job preferences, the career choice of nurse(s) and health worker(s), rural retention. The databases were searched for articles, publications, and dissertations published in English between 2000 and 2021. The major goal of this review was to compile a list of attributes and levels of healthcare workers’ job preferences that had previously been utilized in related studies, as the potential attributes may be employed in a DCE study. The order and frequency of attributes were examined in the study. The checklist for conjoint analysis applications in health care was used to assess the quality of the investigations (31). In addition, semi-structured, in-depth interviews were conducted with seven current nurses and experts in the field of nursing with at least 2 years of working experience. The interview’ s design was based on the instructions proposed by Guion et al (32). Moreover, They have worked in both the public and private sectors and have had experience working in a variety of medical settings. Interviews were performed in a conversational style, allowing participants to freely discuss important subjects. They were questioned indirectly regarding the most important factors that affect nurses’ employment decisions and what work-related circumstances increase nurses’ job satisfaction for working in less desirable situations. Asking indirect questions about attributes allows the experts to express their opinions fully (21). Thus, no attributes were mentioned to experts to avoid any bias of all kinds. Furthermore, they were also questioned about potential incentives that might be provided to persuade nurses to accept or continue serving in rural or remote areas. Interviews were conducted in March 2022. The Skype platform was used for interviews, which provided online conversations. The Persian-language interviews, which lasted between 30 and 45 minutes, were recorded.

#### Stage 2: data reduction

After collecting raw data, the next step is to reduce this data to a limited number of attributes (27). The 68 attributes from the literature that were obtained had some iteration, thus those that belonged to the same group were put into one category after being assessed. Consequently, a candidate list of 23 attributes was acquired In the second stage (consisting of both literature and interview-acquired attributes). Table 2 shows all extracted attributes.

#### Stage 3: removing inappropriate attributes

A DCE with too many attributes makes the responders’ tasks more difficult, which in turn increases error variance, attribute non-attendance (a phenomenon where not all attributes are considered in reaching a decision), and inconsistent responses across choice tasks (28,33–35). Consequently, due to the limitations in the number of attributes and levels that were included in the final design of the DCE, 19 nurses were asked to rate and prioritize the attributes and levels according to their importance. This allows the research team to select the most appropriate attributes and levels for the DCE. Out of a potential 23 attributes, nurses were asked to choose ten and rank them in terms of significance. On a scale of 1 to 10, each attribute was scored. The first priority receives 10 points, while the subsequent selections receive points in decreasing order until the last selection receives one. Attributes that were not among the nurse’ s top 10 priorities were not given any points. To determine an overall score and rank, the scores for each attribute were summed up. Besides, nurses were also asked to use a checklist with 2 to 4 options (levels) for attributes to further prioritize the attribute levels. For each attribute, they were asked to select the best options (levels). The level selection process followed the same steps as the attribute selection step, and the priority of the levels was determined by summing up the responses for each level.

Based on the objectives of the study and experience from previous works, the research team further synthesized and summarized the final levels of attributes. Moreover, there were several criteria applied, including salience, plausibility, the capability of being traded, and relevance to study objectives and decision context (27,28,31,36).

#### Stage 4: attribute wording

The final stage in the process of attribute development aims to ensure that the desired meaning is evoked and that the terminology is understandable for respondents (27). JMP Pro 16 (JMP^®^ Pro *16*. SAS Institute Inc., Cary, NC, 1989–2021) was used to construct the final design of choice sets. It generated a utility-neutral design by setting prior means to 0. In a utility-neutral design, all choices within a choice set are equally probable (37). The design was blocked into 3 versions, each consisting of 6 scenarios (choice sets). Each scenario consisted of 2 profiles. There are six different methods to test the internal validity of DCEs (38). The within-set dominant profile was used as an internal validity test (internal consistency) in the pilot study. Thus, one of the choice sets in each version was included with one dominant profile that was superior to its alternative as a consistency test (21,23). Moreover, the dominant choice set isn’ t considered in the data analysis in the final study. In the end, there were 7 choice sets in each version of the questionnaire. Moreover, the design didn’ t include a no-choice alternative. Finally, socioeconomic and demographic questions were added to the questionnaire. The final questionnaire was piloted on 24 nurses. Each of the eight respondents was required to respond randomly to one of three versions.

## Results

### The scoping review of the literature

A total of 11 papers that were extremely pertinent to the topic of our study were found after the search and review of the inclusion criteria. Table 1 shows a summary of the included studies from the scoping review. Also, it shows that some attributes, such as salary, career development, and workplace facilities, are repetitive across studies. They are, however, expressed in different ways and some modifications have been made to them following the terms of the country of study in order to complete the research appropriately. In these studies, multiple attributes were utilized, which were examined and described comprehensively in the scoping review table. Different systematic methods for obtaining attributes have been reported in various studies. Accordingly, the most common ways of obtaining the candidate list of attributes are by conducting literature reviews, interviewing experts, and focus group discussions. Other studies, except two (17,39), used more than one method to create appropriate attributes. A list of 68 attributes was provided by these studies. Then, these attributes were divided into 21 separate categories. The minimum number of attributes used in these studies was 5 (8,39–42) and the maximum number of attributes was 8 (14,43). Other studies used 6 (44) or 7 (17,22,45) attributes. Attributes that were present in more than 50% of articles [n > 5] are salary [11], workplace facilities [9], lodging [8], location of the facility [7], and workload [6]. Some attributes such as Accessibility and connectivity of workplace location (44), Organizational culture (45), Proximity to family (17), job assignment in an urban area (44) Points when applying for training in family and community health (14), recognition of rural service (14), the sector of the facility (43), benefits (43), financial settlements (22), and work schedule (14) were present in the studies once.

**Table 1.** A summary of the included studies from the scoping review

| Row | Author(s)<br>Year of<br>publication | Study title | Cadre<br>& Sample<br>size | Country<br>of study | The method for<br>selecting<br>attributes | Attributes and levels |
| --- | --- | --- | --- | --- | --- | --- |
| 1 | Bo-hyun Park,<br>YuKyung Ko<br>(2016)<br>(45) | Hospital<br>preferences of<br>nursing students in<br>Korea: a discrete<br>choice experiment<br>approach | Nurses,<br>702 nursing<br>students | Republic<br>of Korea | literature review, in-depth interview | Location [Capital area or large city/ Medium or small<br>city/ Rural or residence area]<br>Hospital type [Tertiary hospitals/ General hospitals]<br>Salary per year [25,000,000 KRW/ 35,000,000 KRW/<br>45,000,000 KRW]<br>Provision of dormitory [Yes/No]<br>Opportunity to upgrade qualifications [3~5 years/ More<br>than 5 years]<br>Welfare system [Sufficient/ Insufficient]<br>Organizational culture [Hierarchy-oriented/ Relations-<br>oriented] |
| 2 | Robyn PJ, Shroff<br>Z, Zang OR,<br>Kingue S,<br>Djienouassi S,<br>Kouontchou C,<br>Sorgho G.<br>(2015)<br>(44) | Addressing health<br>workforce<br>distribution<br>concerns: a<br>discrete choice<br>experiment to<br>develop rural<br>retention<br>strategies in<br>Cameroon | 351 medical<br>students,<br>nursing<br>students, and<br>health workers | Cameroon | literature review,<br>focus group discussion | Accessibility and connectivity of the workplace to the<br>city [Your facility is located in a village with poor<br>connectivity - reliable transportation to the health<br>district capital twice a week or less/ Your facility is<br>located in a village with good connectivity - reliable<br>transportation to the health district capital every day]<br>Health facility infrastructure [Lack of equipment, drugs,<br>and frequent shortages of inputs for most services that<br>you are supposed to provide at the primary healthcare<br>level (delivery, vaccination, prenatal consultation, and<br>general consultation)/ Adequate equipment, drugs, and<br>rare shortages of inputs for most services that you are<br>supposed to provide at the primary healthcare level<br>(delivery, vaccination, prenatal consultation, and general<br>consultation)]<br>Lodging [No accommodation/ A good quality house is<br>made available in a secure location with access to<br>drinking water]<br>Career development [No preferential admission/<br>Establishment of preferential admission for ongoing<br>training available to your level via a quota of 20% of<br>seats reserved for those who worked for at least 4 years<br>in rural areas]<br>Salary (inc. all bonuses) [Your base salary at its current<br>level/base salary + 25% rural retention bonus/ base<br>salary + 50% rural retention bonus/ base salary + 75%<br>rural retention bonus]<br>Job assignment in an urban area [Uncertain/ Automatic<br>after 3 years] |
| 3 | Lindsay J.<br>Mangham and<br>Kara Hanson<br>(2008)<br>(39) | Employment<br>preferences of<br>public sector<br>nurses in Malawi:<br>results from a<br>discrete choice<br>experiment | 232 registered<br>nurses | Malawi | in-depth interview | Place of work [City/District town]<br>Net monthly pay [K30 000 (approximately \$240)/ K40<br>000 (approximately \$320)/ K50 000 (approximately<br>\$400)<br>Availability of material resources [Usually inadequate<br>supply/ Usually adequate supply] |
|  |  |  |  |  |  | Typical daily workload [Light: more than enough time to complete duties/ Medium: enough time to complete duties/ Heavy: barely enough time to complete duties]<br>Provision of government housing [No government housing provided/ Basic government housing provided/ Superior government housing provided]<br>Opportunity to upgrade qualifications [After 3 years/ After 5 years] |
| 4 | Liu S, Li S, Yang R, Liu T, Chen G. (2018) (40) | Job preferences for medical students in China<br>A discrete choice experiment | 519 medical students | China | literature review, in-depth interview, focus group discussion | Location [Township or rural/ County/City]<br>Hospital type [Primary hospital/ Secondary hospital/ Tertiary hospital]<br>Monthly income [3000 CNY/ 6000 CNY/ 9000 CNY]<br>Bianzhi [None/Offer]<br>Training and career development opportunity [Insufficient/Average/ Sufficient]<br>Work environment [Poor/Common/ Superior] |
| 5 | Liu T, Li S, Yang R, Liu S, Chen G (2019) (41) | Job preferences of undergraduate nursing students in eastern China: a discrete choice experiment | 445 undergraduate nursing students | China | literature review, in-depth interview, focus group discussion | Location [Township or village/ County/City]<br>Monthly income [RMB 2000/ RMB 5000/ RMB 8000]<br>Bianzhi [Yes/No]<br>Career development and training opportunity [Insufficient/Some/Sufficient]<br>Work environment [Poor: insufficient essential equipment, limited support by managers, poor superior-subordinate relationship and colleagueship/ Normal: nearly sufficient essential equipment, partly support by managers, moderate superior-subordinate relationship and colleagueship/ Excellent: sufficient essential equipment, full support by managers, harmonious superior-subordinate relationship and colleagueship.]<br>Working strength [Heavy: barely enough time to complete duties, working overtime or night shift four times a week at least/ Medium: nearly enough time to complete duties, working overtime or night shift twice or three times a week/ Light: enough time to complete duties, working overtime or night shift once a week at most] |
| 6 | Rafiei S, Arab M, Rashidian A, Mahmoudi M, Rahimi-Movaghar V. (2015) (17) | Policy interventions to improve rural retention among neurosurgeons in Iran: A discrete choice experiment | 120 neurosurgeons | Iran | in-depth interview | Location [Rural/Urban]<br>Income [Base income/Base + 100% increase/ Base + 150% increase/ Base + 200%]<br>Dual practice [Yes/No]<br>Workload [Light/Moderate/heavy]<br>Proximity to family [Yes/No]<br>Educational facilities [Basic/Superior]<br>Clinical infrastructure [Adequate/Inadequate]<br>Housing [None/Basic/Superior] |
| 7 | Huicho L, Miranda JJ, Diez-Canseco F, Lema C, Lescano AG, Lagarde M, Blaauw D. (2012) (14) | Job preferences of nurses and midwives for taking up a rural job in Peru: a discrete choice experiment | 80 nurses and midwives | Peru | literature review, in-depth interview, focus group discussion | Health facility [Health post (rural)/Health center (rural)/ Health center (city)/ Regional hospital (city)]<br>Monthly take-home (after tax) salary [1000 PEN (rural)/1250 PEN (rural)/1500 PEN (rural)/1750 PEN (rural)/1000 PEN (city)]<br>Time in the post before getting a permanent job [3 years (rural)/6 years (rural)/6 years (city)/10 years (city)]<br>Points when applying for training in Family and Community Health Specialization, after 3 years in post [10 points bonus when applying for training in Family and Community Health Specialization (rural)/ 20 points bonus when applying for training in Family and Community Health Specialization (rural)/none (city)/ 10 points bonus when applying for training in Family and Community Health Specialization (city)]<br>Scholarship for training in Family and Community Health Specialization, after 3 years in post [No (rural)/Yes (rural)/No (city)] |
|  |  |  |  |  |  | Free housing provided [A shared room in a residence with shared facilities (rural)/ A 2-bedroomed independent house (rural)/None (city)<br>Work Schedule (excluding holidays) [You work 22 days and then have 8 days off (rural)/ You work 18 days and then have 12 days off (rural)/ You work every day except Sundays (city)<br>Recognition of rural service [No (rural)/ You get an official certificate of recognition (rural)/No (city)] |
| 8 | Mumbauer A, Strauss M, George G, Ngwepe P, Bezuidenhout C, de Vos L, Medina-Marino A. (2021) (43) | Employment preferences of healthcare workers in South Africa: Findings from a discrete choice experiment | 855 South African healthcare workers | Republic of South Africa | literature review, in-depth interview | Workload [Manageable/Heavy]<br>Workplace Culture [Enabling(“Good”) culture/ Poor culture]<br>Availability of Equipment [Sufficient/Insufficient]<br>Opportunities for Training [Frequent/Infrequent (minimal)]<br>Sector and Facility type [Public clinic/ Public Hospital/ Private clinic/ Private Hospital]<br>Location [Urban; No relocation/ Urban; Relocation/ Rural; No relocation/ Rural; Relocation]<br>Salary [5% increase on current salary/ 10% increase on current salary/ 20% increase on current salary/ Salary cut of 20%]<br>Benefits [None/ Medical aid contribution to the value of 7% of the salary/ Medical aid and pension contribution of 20% of the salary/ Medical aid, pension, and housing contribution of 40% of the salary] |
| 9 | Rockers PC, Jaskiewicz W, Wurts L, Kruk ME, Mgomella GS, Ntalazi F, Tulenko K. (2012) (8) | Preferences for working in rural clinics among trainee health professionals in Uganda: a discrete choice experiment | 132 nursing students | Uganda | literature review, in-depth interview, focus group discussion | Salary [450,000 USh per month/ 550,000 USh per month/ 650,000 USh per month/ 750,000 USh per month]<br>Facility quality [Basic/Advances]<br>Housing [No housing or allowance provided/ Housing allowance provided, enough to afford basic housing/ Free basic housing provided]<br>Length of commitment [2 years/5 years]<br>Support from manager [The district health officer is not supportive and makes work more difficult/ The facility manager is supportive and makes work easier]<br>Facility staffing [“You have extra responsibility because 50% of health worker positions are vacant/ You have extra responsibility because 25% of health worker positions are vacant/ You don’t have extra responsibility because all health worker positions are filled] |
| 10 | Kazemi Karyani A, Karami Matin B, Malekian P, Moradi Rotvandi D, Amini S, Delavari S, Soltani S, Rezaei S. (2021) (22) | Preferences of Medical Sciences Students for Work Contracts in Deprived Areas of Iran: A Discrete Choice Experiment Analysis | 201 medical students | Iran | literature review, interview | Payment [Capitation/Capitation +30% bonus/ Capitation +50% bonus]<br>Place of medical center [Own province/ A nearby province/ A faraway province]<br>Financial settlements [max to 15 days/ Between 15 to 30 days/ Between 30 to 60 days]<br>Housing and transportation facilities [No/Yes]<br>Duration of contract [One year/3 years/5 years]<br>Quotations for continuing education [No/ After 4 years’ work in deprived area/ After 8 years’ work in deprived area]<br>Workload [Low/Moderate/Heavy] |
| 11 | Prust ML, Kamanga A, Ngosa L, McKay C, Muzongwe CM, Mukubani MT, Chihinga R, Misapa R, van den Broek JW, Wilmink N. | Assessment of interventions to attract and retain health workers in rural Zambia: a discrete choice experiment | 474 practicing health workers and students | Zambia | in-depth interviews, focus group discussion | Salary [Base salary/ Base salary, plus 20% rural allowance/ Base salary, plus 25% rural allowance/ Base salary, plus 30% rural allowance]<br>Educational opportunities [No scholarship after 2 years: guaranteed paid leave after 2 years with no government financial assistance for study/ 75% scholarship after 3 years: guaranteed paid leave after 3 years with eligibility for (not guaranteed) 75% government scholarship for study/ 100% scholarship after 4 years: guaranteed paid |
|  | (2019) (42) |  |  |  |  | leave after 4 years with 100% government scholarship for study guaranteed]<br>Housing [Basic allowance offered (20% of base salary), but no housing provided/ Basic housing provided (two bedrooms, outside bathroom, no electricity, water available through a borehole or hand pump)/ Superior housing provided (three bedrooms, electricity, piped running water, self-contained master bedroom and security reinforcements)/ Superior allowance offered (30% of base salary), but no housing provided]<br>Transportation [None/ Available: reliable access to ambulance and utility vehicle (motorbike/vehicle) for official facility use only]<br>Medical equipment [Inadequate: a standard list of medical equipment at health facility not always available/ Adequate: a standard list of medical equipment at health facility always available] |

### Interview and rating

Interviews with seven nurses and nursing experts were conducted. They all agreed that the most critical element influencing nurses’ employment decisions was salary. The interviewees believed that although salary is the most critical factor in rural retention policies, policymakers can offer a package consisting of monetary and non-monetary incentives to healthcare workers that can reduce the financial burden of such policies on the government. The interviewees’ suggestions were close to the attributes that have been previously used in the literature. However, in accordance with the working conditions and job hierarchy in Iran, two new attributes— the expected time spent on the assigned job for promotion to a higher position and the type of employment contract—were subsequently created. Salary, location of the health facility, and workload were the top three attributes that received the highest scores, respectively. The remaining seven important attributes were work schedule, type of health facility, type of employment contract, workplace facilities, expected time spent on the assigned job for promotion to a higher position, lodging, and workplace culture. Researchers reviewed all of the attributes and levels obtained from the data from earlier stages. Finally, with consideration of the exclusion (stage 3) and objectives of the study, salary, type of employment contract, workload, type of health facility, workplace facilities, work schedule, and expected time spent on the assigned job for promotion to a higher position were seven attributes considered in the final design of the scenarios. Table 2 shows all extracted attributes.

**Table 2.** Extracted attributes from the literature review and interviews

| Row | Final Attributes | Number of presence in the studies | Source of attribute |  | Is it a part of the final design? | Levels |
| --- | --- | --- | --- | --- | --- | --- |
|  |  |  | Literature | Interview |  |  |
| 1 | Location of the facility | 7 | * | * |  |  |
| 2 | Type of health facility | 4 | * | * | y | Public hospital/Private hospital/Clinic/Health house (Health post) |
| 3 | Salary | 11 | * | * | y | 90 million IRR/110 million IRR/130 million IRR (as monthly wage) |
| 4 | Lodging | 8 | * | * |  |  |
| 5 | Career development | 4 | * | * |  |  |
| 6 | workplace facilities | 9 | * | * | y | Poor/Normal/Excellent |
| 7 | Organizational culture | 1 | * |  |  |  |
| 8 | Accessibility and connectivity of workplace location | 1 | * |  |  |  |
| 9 | Job assignment in an urban area | 1 | * |  |  |  |
| 10 | Workload | 6 | * | * | y | Low/Moderate/Heavy |
| 11 | Workplace culture | 3 | * | * |  |  |
| 12 | Proximity to family | 1 | * |  |  |  |
| 13 | Educational opportunities | 3 | * |  |  |  |
| 14 | Length of commitment | 3 | * | * |  |  |
| 15 | Points when applying for training in Family and Community Health Specialization, after 3 years in post | 1 | * |  |  |  |
| 16 | Recognition of rural service | 1 | * |  |  |  |
| 17 | The sector of the Facility (public/private) | 1 | * | * |  |  |
| 18 | Type of employment contract | - |  | * | y | Gharadadi/Rasmi/Peymani |
| 19 | Benefits | 1 | * |  |  |  |
| 20 | Financial settlements | 1 | * |  |  |  |
| 21 | Transportation | 2 | * |  |  |  |
| 22 | Expected time spent on the assigned job for promotion to a higher position | - |  | * | y | 1 to 3 years must be spent on the assigned job promotion/3 to 5 years must be spent on the assigned job promotion/More than 5 years must be spent on the assigned job promotion |
| 23 | Work Schedule | 1 | * | * | y | Appropriate/Inappropriate |

### Pilot study

The pilot study was conducted to test its level of complexity as well as the validity and reliability of the data. A utility-neutral design was constructed by JMP. Internal and face validity, intelligibility, and acceptability of the questionnaire were tested in the pilot study, and minor modifications were made. The pilot study is used to determine the clarity and appropriateness of the choice sets (21,23). It was revealed that the respondents in the pilot study were able to understand and answer all of the choice sets with ease. Additionally, all of the respondents successfully passed the consistency test. The questionnaire took an average of 6 minutes to complete. The result of the pilot study would be used to generate a local D-optimal design for conducting a DCE on nurses’ job preferences. The local D-optimal design takes into account the prior of the mean, but does not include any information from a prior covariance matrix (37). JMP was also used to analyze the data of the pilot study and the results are shown in tables 3, 4, and 5.

**Table 3.** The results of the logit model and willingness to pay for job attributes of the pilot study

| Attributes and their levels | $\beta$ | SE | Chi square | P-value | 95% CI | WTP | 95% CI |
| --- | --- | --- | --- | --- | --- | --- | --- |
| Monthly salary (unit: 10 000 000 IRR) | 1.458 | 0.385 | 24.340 | <0.001 | 0.694 2.885 |  |  |
| Type of employment contract (ref: Rasmi) |  |  | 16.004 | <0.001 |  |  |  |
| Gharardadi | -0.450 | 0.332 |  |  | --1.609 0.262 | 1.249 | 0.0001 2.498 |
| Peymani<sup>1</sup> | -0.921 | 0.325 |  |  | -2.128 -0.347 | 1.571 | 0.372 2.770 |
| Workload (ref: heavy) |  |  | 18.169 | <0.001 |  |  |  |
| Low | 0.849 | 0.309 |  |  | 0.260 1.911 | -1.433 | -2.466 -0.401 |
| Moderate | 0.393 | 0.256 |  |  | -0.142 1.150 | -1.121 | -1.879 -0.363 |
| Type of facility (ref: health house) |  |  | 2.598 | 0.457 |  |  |  |
| Public hospital | 0.764 | 0.422 |  |  | -0.260 2.101 | -1.194 | -2.068 -0.321 |
| Private hospital | 0.393 | 0.348 |  |  | -0.505 1.271 | -0.940 | -1.840 -0.039 |
| Clinic | -0.179 | 0.354 |  |  | -1.019 1.066 | -0.547 | -1.403 0.307 |
| workplace facilities (ref: excellent) |  |  | 7.921 | 0.003 |  |  |  |
| Poor | -0.739 | 0.288 |  |  | -1.523 -0.201 | 0.804 | 0.112 1.495 |
| Normal | 0.306 | 0.272 |  |  | -0.419 0.966 | 0.086 | -0.538 0.711 |
| Work schedule (ref: appropriate) |  |  | 8.743 | 0.003 |  |  |  |
| Inappropriate | -0.836 | 0.325 |  |  | -2.372 -0.249 | 1.147 | -0.046 2.340 |
| Expected time spent on the assigned job for promotion to a higher position (ref: More than 5 years) |  |  | 0.101 | 0.95 |  |  |  |
| 1 to 3 years | 0.461 | 0.364 |  |  | -0.366 1.482 | -0.764 | -1.757 0.228 |
| 3 to 5 years | 0.191 | 0.323 |  |  | -0.754 1.381 | -0.579 | -1.477 0.319 |
| AICc | 113.489 |  |  |  |  |  |  |
| BIC | 149.297 |  |  |  |  |  |  |
| -2Log Likelihood | 84.689 |  |  |  |  |  |  |
| -2Firth Log Likelihood | 50.084 |  |  |  |  |  |  |

SE standard error, WTP willingness to pay, CI confidence interval, AICc Akaike information criterion (for small sample size), BIC Bayesian information criterion

**Table 4.**
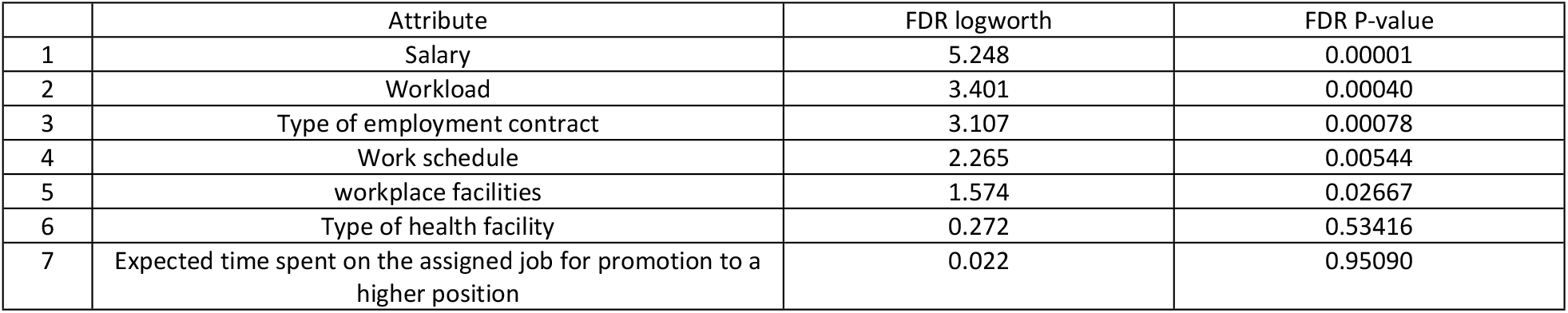
The significance of attributes

### Statistical analysis

Random utility theory provides the theoretical foundation for analysis of the DCEs data. The utility (U) associated with a particular job is made up of 2 components: the deterministic component V_ni_ (where V is a function of observable characteristics) and the unobservable component ε_ni_ (23,40). The utility, U, to individual n associated with job i can be specified as

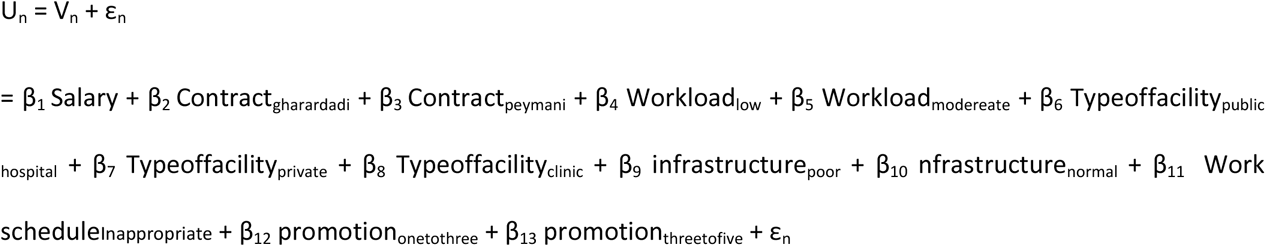

The beta (β) coefficients generated from the logit model in the equation can be used for two main purposes (46):

- To determine whether Attributes are statistically significant.
- The direction of the signs of the coefficients also provides an examination of the theoretical validity of the DCE model, that is, whether the coefficients move according to economic theory or predict expectations or not.

In the choice platform of JMP, the parameter estimates report gives estimates and standard errors of the coefficients of utility associated with the effects (attributes) listed in the term column of the platform (47). Table 3 shows the result of the conditional logit model.

As it’ s shown in table 4, salary is the most significant factor that affects nurses’ job-selecting decisions. In addition, non-monetary attributes are also important. Table 5 shows the significance of attributes using false Discovery Rate LogWorth for each model effect, defined as -log10(FDR PValue). This is one of the best statistics for plotting and assessing significance (48). Willingness to pay is the amount of money that a nurse expects to be paid more for accepting or withdrawing from a level to another level of an attribute. For example, nurses were willing to be paid 11 147 000 IRR more to accept a job with an inappropriate work schedule.

## Discussion

A discrete choice experiment (DCE) is a quantitative technique for eliciting individual preferences (29). By calculating the marginal rate of substitution between different influencing factors on these preferences, DCE enables researchers to investigate the trends in the trade-offs (45). As DCEs are increasingly popular to value outcomes for health economic studies and gradually gain acceptance as an input into policy decisions, results may misguide decision-makers if they are based on an inappropriate set of attributes (27). Furthermore, health-related DCEs rarely comprehensively conduct and report the attribute and level selection process (28,49). Nevertheless, some researchers have proposed systematic methods to develop attributes for DCEs in detail (27,28). Therefore, we aimed to establish attributes and levels for a discrete choice experiment on nurses’ job preferences and transparently report the whole process. Regarding the framework of Helter and Boehler, this study was conducted in four stages. For the first two stages, we used a combination of two methods: literature review and qualitative methods. Using literature reviews alone may lead to the omission of some relevant attributes which may, in turn, increase the error variances and introduce bias into the study (50,51). Therefore, qualitative studies are advocated for as they help in identifying context-specific attributes that are important to the study respondents (26,51,52). Furthermore, qualitative studies can also help in revealing new attributes not captured in the literature (28). Several studies have adopted such strategies (52,53). Hence, we performed a scoping literature review and in-depth interviews to identify the factors influencing nurses’ job preferences and to understand the importance of each attribute for nurses in the first two stages. These stages resulted in two new attributes: type of employment contract and expected time spent on the assigned job for promotion to a higher position. In the third stage (removing inappropriate attributes), by the end of these stages, 23 potential job-related factors that affect nurses’ career choices were identified. Finally, based on the acquired information and qualitative data, the research team selected the final attributes and levels. In the fourth stage, a pilot study was conducted. In this document, the pilot study’ s findings were also reported. Willingness to pay was calculated and shown in Table 3. Table 2 lists the final attributes and levels along with the attributes that were taken from the interviews and literature reviews.

### Strengths and limitations

This study has several strengths. First, this study was among the first attempts to address the rural retention of health workers (nursing cadre) in Iran. Second, the process of establishing attributes used a systematic method and was reported in detail. This can lead to an improvement in transparency. Third, in contrast with other studies (21,40,41), the data from the pilot study was also analyzed and the results were reported rigorously.

On the contrary, there were some limitations in this study. First, in order to shorten the questionnaire for conducting online, considering the previous experiences of the research team with online questionnaires, it was necessary to make it as brief as possible. Another reason was to prevent discomfort and refusal to continue the questionnaire by the respondents. Due to these reasons, a simple method of testing the internal validity (within-set dominated pairs) of the questionnaire was used. It increased the total number of choice sets to 7 in each block, while using other more complicated methods like the transitivity test would have unnecessarily increased the number of choice sets to 9. Second, the sample size of the pilot study might have been too small. This might be the reason why two attributes were not statistically significant (P >0.05).

## Conclusion

Some of the most significant financial and non-financial factors that affect Iranian nurses’ job preferences are reported in this study. They were gathered through a review of the literature or interviews with experts. This indicates that policymakers have a wide range of interventions available that can significantly improve the working conditions of nurses. In addition, the full description of the attribute development and level selection processes makes this study a valuable contribution to the literature on DCE.

## Data Availability

All relevant data are within the manuscript and its Supporting Information files.

## Acknowledgments

This paper was part of the Master thesis of Amirmohammad Haddadfar and received the ethical approval of the Research Ethics Committees of the School of Public Health & Allied Medical Sciences-Tehran University of Medical Sciences (approval ID: IR.TUMS.SPH.REC.1400.344). All participants also completed an informed consent form before being interviewed or completing the pilot DCE survey questionnaire.

## Supporting information

S1 file: Interview (pdf)

S2 file: Questionnaire (pdf)

## Footnotes

1 According to the national employment law of Iran, Rasmi is a kind of employment contract between two parties (mostly the government and its employees) that includes more benefits, such as job security, than other employment contracts. Peymani and Gharardadi differ in terms of the contract length and other benefits.

